# The role of children in the transmission chain of SARS-CoV-2: a systematic review and update of current evidence

**DOI:** 10.1101/2020.11.06.20227264

**Authors:** Jonathan E. Suk, Constantine Vardavas, Katerina Nikitara, Revati Phalkey, Jo Leonardi-Bee, Anastasia Pharris, Emma Wiltshire, Tjede Funk, Lisa Ferland, Nick Bundle, Jan C. Semenza

**Author notes:** Corresponding author: Jonathan E. Suk. Gustav III:s Boulevard, 169 73 Solna, Sweden. The authors contributed equally to this manuscript (Jonathan E. Suk, Constantine Vardavas).

## Abstract

Decisions on school closures and on safe schooling during the COVID-19 pandemic should be evidence-based. We conducted a systematic literature review to assess child-to-child and child-to-adult SARS-CoV-2 transmission and to characterise the potential role of school closures on community transmission. 1337 peer-reviewed articles published through August 31, 2020 were screened; 22 were included in this review. The literature appraised provides sufficient evidence that children can both be infected by and transmit SARS-CoV-2 in community, household and school settings. Transmission by children was most frequently documented in household settings, while examples of children as index cases in school settings were rare. Included studies suggested that school closures may help to reduce SARS-CoV-2 transmission, but the societal, economic, and educational impacts of prolonged school closures must be considered. In-school mitigation measures, alongside continuous surveillance and assessment of emerging evidence, will promote the protection and educational attainment of students and support the educational workforce.

## Introduction

One of the more perplexing and controversial dimensions of the COVID-19 pandemic surrounds the role of children in the transmission. Are they drivers of the pandemic, or are they merely innocent bystanders, affected in myriad ways by school closures and other physical distancing measures while not being generally at-risk of COVID-19 themselves?

During the first few months of the COVID-19 pandemic, the vast uncertainty surrounding the epidemiology of SARS-CoV-2 led many countries globally to include school closures as part of more general lockdowns. In the European Union (EU) / European Economic Area (EEA), all but two countries, Iceland and Sweden closed primary schools roughly during the period of March 2020 – May 2020 **(Figure 1**); all EU/EEA countries closed secondary schools, to some degree, during the same period **(1)**. These school closures were concomitant with other societal-level physical distancing measures during the first wave of the COVID-19 pandemic. Available data from The European Surveillance System (TESSy) indicates that severe outcomes from SARS-CoV-2 infections, including fatalities, are much lower in paediatric patients compared to adults (**Figure 2**). As of 1 November 2020, and from over a million reported COVID-19 cases in Europe from 12 countries (Cyprus, Estonia, Finland, Germany, Ireland, Italy, Latvia, Lithuania, Luxembourg, Norway, Poland and Slovakia), rates of hospitalization and severe hospitalization (i.e. intensive care unit admissions, respiratory support or oxygen therapy) are notably lower for children than for adults (particularly for those over 40 years of age), with one notable exception being hospitalization rates for infants under 1 year (**Figure 2**). Meanwhile, fatalities among children due to COVID-19 are very rare: as of 1 November 2020, across these 12 countries in the European Union and European Economic Area (EU/EEA), there have been 12 reported fatalities for children under 16 years of age (of 67,701 reported cases), and six deaths for children between 16 and 19 years of age (of 42,545 reported cases). Since September 2020, which coincides with start of the new school year, the age-specific risk of severe outcomes has fallen in all age groups (**Figure 2, panels a-c**).

**Figure 1.**
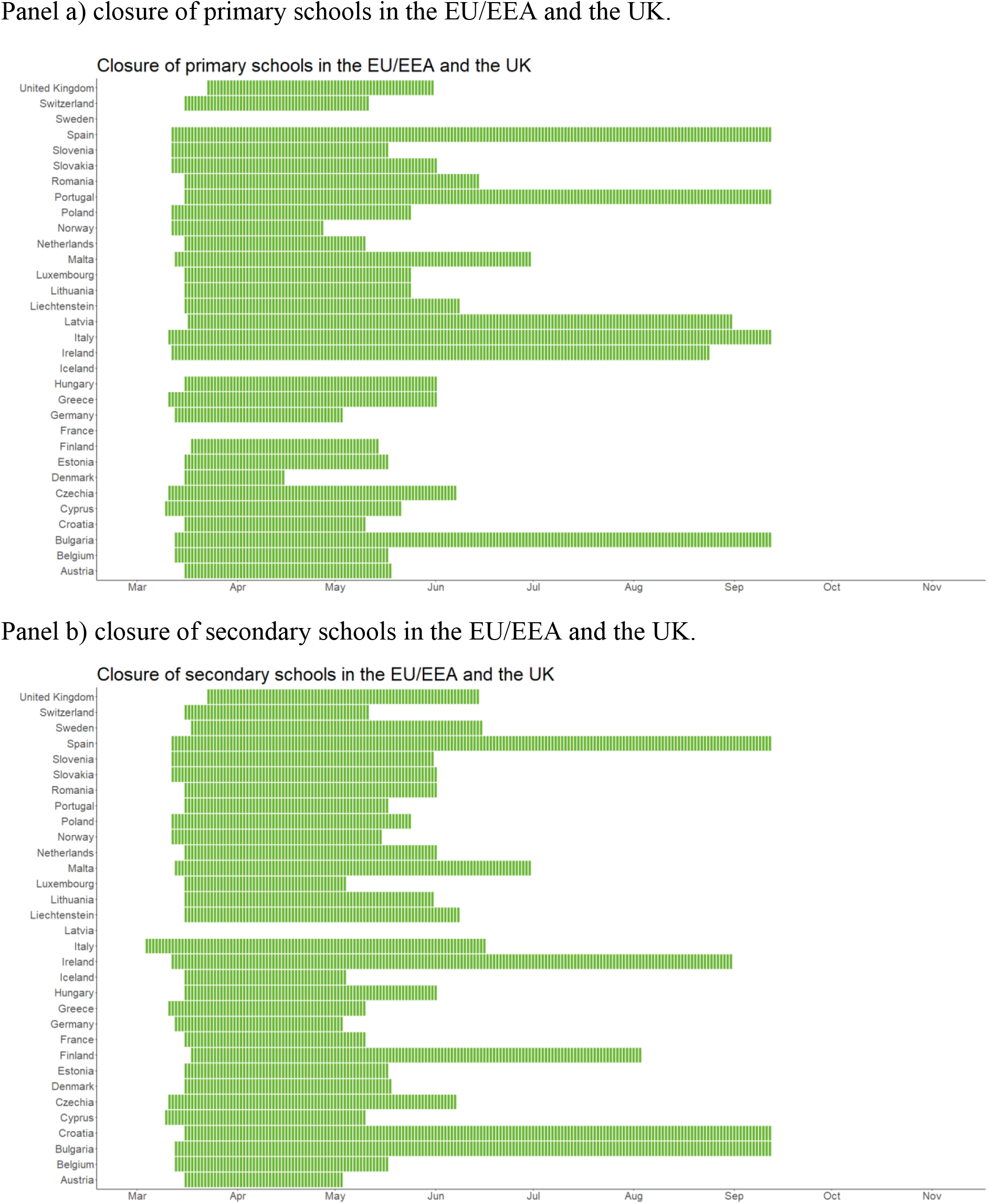
Timing of the closure of primary (panel a) and secondary (panel b) schools in the EU/EEA and the UK March-September 2020 Panel a) closure of primary schools in the EU/EEA and the UK.

**Figure 2.**
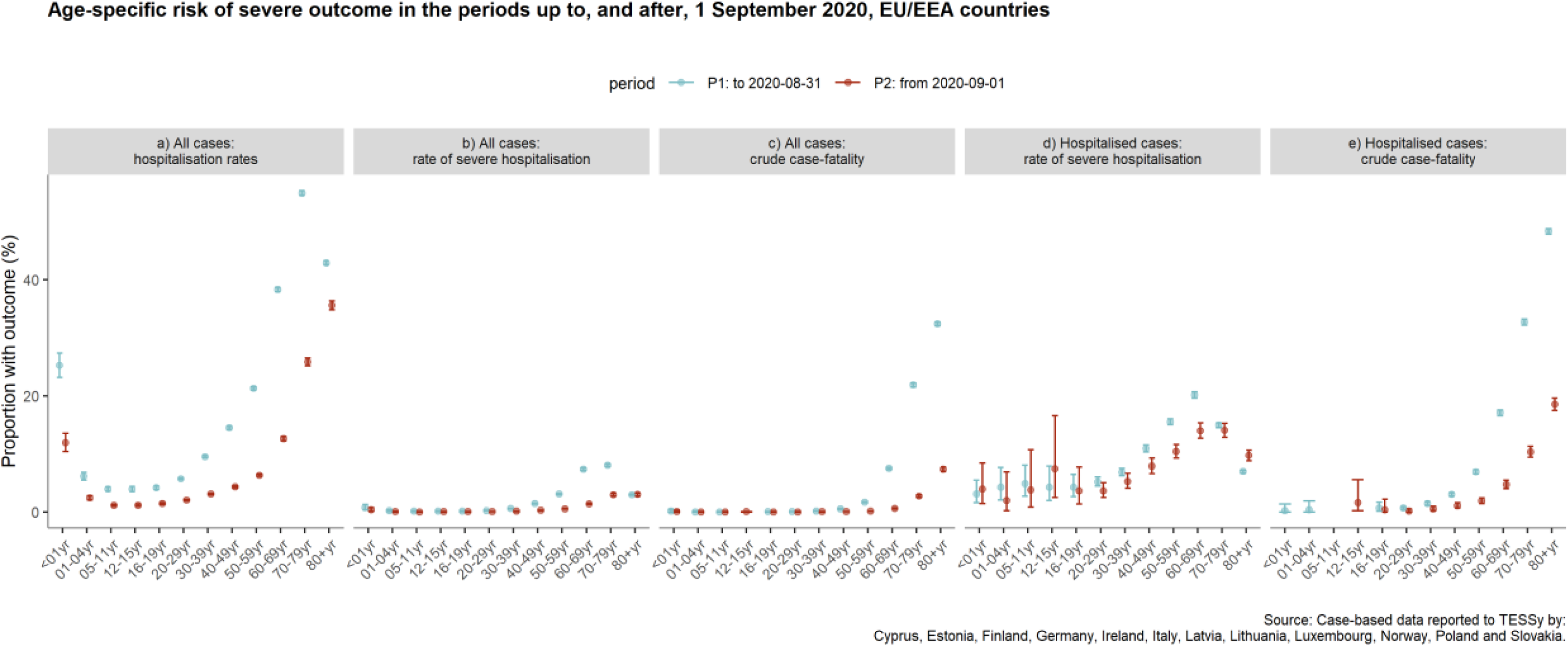
Age-specific proportion of severe outcomes of COVID-19 in the periods up to, and after 1 September 2020, the European Surveillance System (TESSy) (data extracted 4 November, 2020 for the period up to 1 November 2020)

Other epidemiologic indicators of SARS-CoV-2 infection in children provide a complex picture regarding their potential role in the transmission chain. A systematic review concluded that, based on current evidence, children and adolescents have lower susceptibility to SARS-CoV-2 infection (2). However, when infected and symptomatic, children appear to shed viral RNA in similar quantities to adults (3), and that younger children (under 5 years) with mild to moderate symptoms may shed even more virus than older children and adults (4). While the proportion of asymptomatic SARS-CoV-2 infections among children in the general population is uncertain, 16% of paediatric cases in Europe were classified as asymptomatic (5), while up to 90% of paediatric cases in China were deemed to be asymptomatic, mild, or moderate (6). It is possible that children are less often asymptomatic carriers than adults: a study of non-COVID-19-related hospitalizations in Milan identified 1% of children and 9% of adults as asymptomatic carriers of SARS-CoV-2 (7).

Important potential sources of evidence surrounding the role of children in the COVID-19 pandemic come from studies situated in the community, household, or school settings and from modelling studies. As the emerging literature may have important insights to guide decision- making around school closures as well support decision making for mitigation measures in household and school settings, and to complement the literature on the susceptibility of children to SARS-CoV-2 (2), a systematic literature review was conducted to assess the role of children in the transmission of SARS-CoV-2.

The primary aim of this systematic literature review was to assess child-to-child and child-to- adult SARS-CoV-2 transmission and secondarily to characterise the potential role of the opening and closing of schools on community transmission. Here we outline the findings from this review, while highlighting perspectives of the European Centre for Disease Prevention and Control (ECDC) on current uncertainties and research gaps, mitigation measures for schools, community and household settings, and surveillance and monitoring priorities.

## METHODS

### Search Strategy

This systematic literature review is reported in accordance with the Preferred Reporting Items for Systematic Reviews and Meta-Analysis (PRISMA) guidelines (8). The methodology used adhered to PRISMA (Preferred Reporting Items for Systematic Reviews and Meta-Analysis) guidelines. Relevant studies published between December 2019 and August 31, 2020 were identified by searching Medline and Embase. The following set of inclusion criteria were used to determine eligibility of the studies, which is based on the PCC framework (P-Population, C- Concept, C-Context). The study *Population* was restricted to people 1-17 years old (excluding neonatal transmission), the *Concept* was to assess child-to-child and child-to-adult transmission, while the *Context* was to assess community, household and school transmission. Subject heading terms and free text words relating to the Population, Concept and Context terms as identified in the inclusion criteria were used to develop a comprehensive list of terms for the search strategy (Supplementary Table 1). We included all studies of quantitative research, while opinion pieces, commentaries, editorials were excluded. We additionally screened reference lists of the included articles to identify further relevant studies. The search was limited to the English language.

### Study selection

Initially, a pilot screening process was used where 100 identical articles were screened for their eligibility independently by two reviewers to ensure consistency in screening. As a high measure of inter-rater agreement was achieved between the two reviewers during the pilot assessment (percentage agreement >90% and/ or Cohen’s Kappa >0.81), the remaining titles were randomly allocated to the two reviewers and screened for eligibility independently by them. After an initial selection of the titles, each reviewer assessed each other’s selected studies. The retrieved articles were then independently double-screened by two reviewers based on the full text of the articles.

### Data extraction

The data extraction template was piloted independently by the two reviewers on a random sample of two included studies to enable an assessment of consistency in data extraction and to identify where amendments needed to be made to the template. The remaining studies were then data extracted independently by two reviewers, and the results were double checked across the original manuscript by a third reviewer.

### Data synthesis

Characteristics of the included studies were presented in tabulated form detailing the study design, geographical location of the study, sample size, characteristics of the populations considered, setting, context, and the findings of the study. A narrative synthesis approach was applied to look systematically at the data and to describe each study categorized by the study design. Patterns in the data were identified through tabulation of results, and an inductive approach was taken to translate the data to identify areas of commonality between studies. Mathematical modelling and simulation studies were assessed separately from those reporting real-life data.

## Results

### Study selection and description

A total of 1411 studies were identified according to the specified selection criteria from Medline and Embase. After the removal of duplicates, 1337 were screened by title/abstract, out of which 102 were assessed via full text, and 22 studies subsequently included in this review. Overall, 14 of the included studies reported on child-to-adult transmission of SARS-CoV-2, while eight explored the impact of school closures on the epidemiology of the COVID-19 pandemic. Almost all studies (7/8) that assessed the impact of school closure were simulation models, while all 14 studies that assessed child-to-adult transmission patterns were observational in design. The PRISMA flowchart showing the flow of study selection is presented in **Figure 3**.

**Figure 3.**
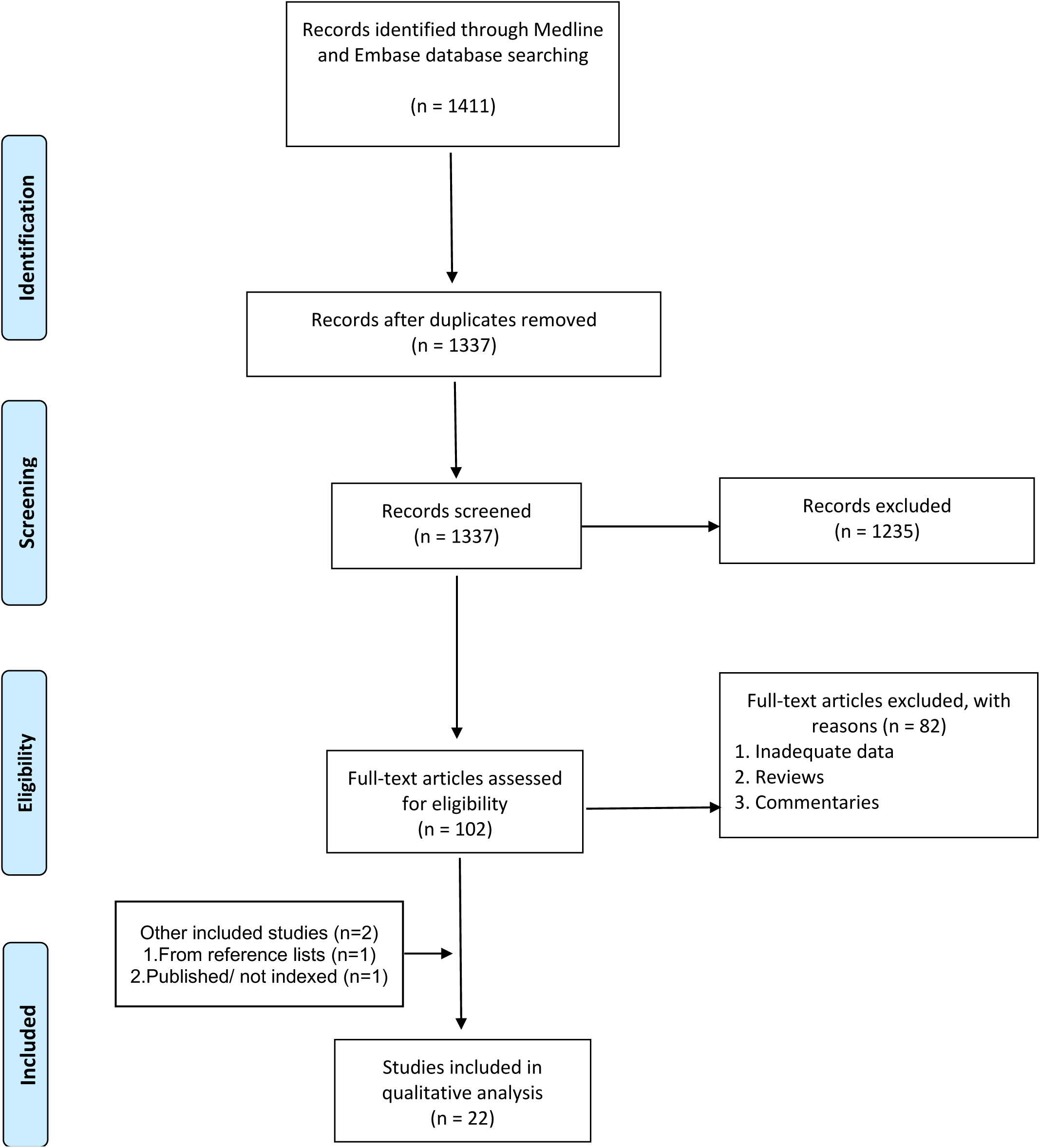
PRISMA Flowchart of study selection included in the rapid review.

### Child-to-child and child-to-adult transmission of SARS-CoV-2

Fourteen published studies were identified to address child-to-adult transmission of SARS-CoV-2. Two referred to transmission within a community setting, six within a household and six within a school setting.

#### Community settings

Only one case study and one ecological study assessed child-to-adult transmission of COVID- 19 within a community setting. A case study by Jung et al. (9), reported the contact-tracing results of a 9-year-old infected asymptomatic girl. A total of 1206 close and casual contacts were identified of which 96% underwent a SARS-CoV-2 PCR test and for which the results were negative for every close and casual contact, except for one adult. Goldstein & Lipsitch (2020) (10) used data on COVID-19 cases in Germany evaluated the contributing role of different age groups in COVID-19 community transmission. According to their analyses, COVID-19 cases among 10-14 and 15-19 year olds were 0.78 times lower and 1.14 times more likely respectively to be a case compared to other age groups after the implementation of physical distancing measures. In comparison to all age groups, individuals aged 10-14 years had lower odds of being a case compared to adults 15-34 years old, while 15-19 year olds were less likely to be a case (OR 0.81) than 20-24 year olds, after the implementation of physical distancing measures, including school closure.

#### Household settings

Four studies assessed household child-to-child and child-to-adult transmission within family clusters as detailed in **Table 1**. Posfay-Barbe et al. assessed the clinical presentation of the 111 household contacts of the first pediatric cases of COVID-19 in Geneva, Switzerland, according to which only in 3/39 (8%) of households the study child was the index case (11). Similarly, Teherani et al., conducted contact tracing among household members of 32 confirmed pediatric COVID-19 cases, for which only in 7/32 households did children develop first symptoms of COVID-19 and were assumed as the suspected index cases (12). Somekh et al. also investigated the role of children in household COVID-19 transmission in Israel, using a cluster of 13 families, and found that only in one family the index case was a child (13). Maltezou et al. noted that among 23 family clusters in Greece only in 2 families the index case was a child, while there was no evidence of child-to-adult or child-to-child secondary transmission (14).

**Table 1.** Studies assessing household child-to-child and child-to-adult transmission within family clusters.

| Author | Country | Population | Timeframe | Number of cases | Median Age of cases (Range or IQR) | Household Contacts | Child index case in household |
| --- | --- | --- | --- | --- | --- | --- | --- |
| Posfay-Barbe et al., 2020 (10) | Switzerland, Geneva | 39 family clusters | 10 March to 10 April, 2020 | 39 | 11.1 (5.7 to 14.5) | 111 | 3/39 |
| Teherani et al., 2020 (11) | USA, Atlanta | 32 family clusters | 16 March to 14 June 2020 | 32 | 12.7 (IQR: 8.3-15.7) | 144 | 7/32 |
| Somekh et al., 2020 (12) | Israel, Bnei Brak | 13 family clusters | n/r | 1 | 0.5-17 | n/r | n/r |
| Maltezou et al., 2020 (13) | Greece | 23 family clusters | 26 February to 3 May 2020 | 1 | 0-18 | 109 | 2/23 |

n/r: not reported

Two studies from South Korea assessed secondary attack rates when children were the index cases as depicted in **Table 2**. Park et al. performed an assessment of 46 COVID-19 index cases (three cases 0-9 years old and 43 cases 10-19 years old) in households. For index cases under 9 years old, the household secondary attack rate was 5.3%, the non-household 1.1% and the overall secondary attack rate was 2.1%. On the contrary, for index cases 10-19 years of age, the secondary attack rates were 18.6% (household), 0.9% (non-household) and 9.8% (overall) (15). An additional study from South Korea among 107 index children and 248 household members, identified only one case of secondary transmission to a household member and the secondary attack rate was estimated to be 0.5%. In this study an additional 40 confirmed cases were found in household members, but it could not be determined as a secondary transmission as they had the same exposure as the pediatric index case (16).

**Table 2.** Studies that assessed the secondary household attack rate, when children are the household index case.

| Author | Country | Population | Timeframe | Number of index cases | Age of cases (Range or IQR) | Positive/traced | Secondary Attack Rate |  |  |
| --- | --- | --- | --- | --- | --- | --- | --- | --- | --- |
|  |  |  |  |  |  |  | Household | Non household | Overall |
| Park et al., 2020 (14) | South Korea | 29 and 124 COVID-19 index cases 0-9 and 10-19 years of age | 20 January to 27 March 2020 | 29<br>124 | 0-9 & 10-19 | 29/237<br>124/457 | 3/57 (5.3%)<br>43/231 (18.6%) | 2/180 (1.1%)<br>2/226 (0.9%) | 5/237 (2.1%)<br>45/457 (9.8%) |
| Kim et al., 2020 (15) | South Korea | 107 pediatric COVID-19 index cases | 20 January to 6 April 2020 | 107 | Median: 15, (IQR 10-17) | 1/248 | 1/248 (0.5%) | n/r | n/r |

n/r: not reported

#### School settings

Six studies assessed SARS-CoV-2 transmission in a school setting the main results of which are outlined in **Table 3**. Heavey et al. conducted a case study in order to explore the role of transmission among children in the school setting in the Republic of Ireland, before school closure (17). Three pediatric and three adult cases of COVID-19 with a history of school attendance were detected with 1155 contacts. Child-to-adult transmission or child-to-child transmission was not reported in this study. Similarly Danis et al. presented the contact tracing results of a nine-year-old child in France, who visited 3 different schools the first days of symptom appearance. There was no evidence of secondary transmission in any of the school contacts (18). Moreover, Yung et al. traced three COVID-19 cases (2 pediatric and 1 adult) in three different educational settings, and the results were negative (19). One study from New South Wales, Australia presented an overview of COVID-19 cases and transmission in schools. In a total number of 25 schools and 10 Early Childhood Educational and Care Settings, 27 index cases were identified, among which 12 were children and 15 staff members. Secondary transmission was noted only in four out of 25 educational settings, and the overall child-to-adult attack rate was estimated at 1.0%. (20).

**Table 3.** Studies assessing SARS-CoV-2 transmission in a school setting.

| Author | Country | Timeframe | Age Range | Setting | Number of index cases | Symptomatic secondary cases in the school settings<sup>1</sup> | Parallel interventions |
| --- | --- | --- | --- | --- | --- | --- | --- |
| Heavey et al., 2020 (16) | Ireland | March 2020 | 10-15 | Schools in Ireland | 6 cases (3 children, 3 staff) | 0/822 child contacts | Exposure before school closure. Schools closed, contacts were quarantined |
| Danis et al., 2020 (17) | France | January to February 2020 | n/a | 3 schools | 1 child | 0/86 in school contacts<br>1/6 hospitalised contacts | n/a |
| Yung et al., 2020 (18) | Singapore | February to March 2020 | 2.8-15 | 3 schools | 3 cases (2 pediatric-1 adult) | 0/42 (symptomatic) | Contacts were quarantined<br>Targeted measures at the school level |
| Macartney et al., 2020 (19) | Australia, NSW | 25 January to 10 April 2020 | <18 | 25 schools and 10 childcare settings | 27 cases (12 children, 15 adults) | 18/1448 overall contacts<br>5/914 in 3 schools<br>(5/18 were asymptomatic) | Contacts were quarantined |
| Stein-Zamir et al., 2020 (20) | Israel | May 2020 | 12-18 | one high school | 153 children and 25 adults | n/a | Closed spaces with poor ventilation, high temperatures, crowded spaces and close contact with no masks. |
| Link-Gelles et al., 2020 (21) | USA, RI | June – July 2020 | <18 | 666 educational settings | 33 confirmed and 19 probable cases in 29 settings | 17 cases in 4/666 educational settings with. | Class distancing, the use of face masks for adults, universal symptom screening daily and disinfection. |

1: Measured from the date of last contact
2: Probable cases

Two studies reported on the regional evidence after the re-opening of schools. A school outbreak in Israel after reopening of schools in May 2020 was described by Stein-Zamir et al. (21). The outbreak assessment was initiated by two pediatric COVID-19 cases that were not epidemiologically related. The results showed that 153/1161 students and 25/151 staff members tested positive for COVID-19. However, this outbreak was attributed to crowded classes, combined with the exemption of facemasks and the use of air-condition due to an extreme heatwave. On the contrary, a study by Link-Gelles et al., in Rhode Island, USA. among 666 child care programs that reopened on 1 June, 2020 after a 3-month closure revealed 52 confirmed and probable cases (33 confirmed cases), of which 30 were among children and 22 among adults (22). Secondary transmission for 10 cases was noted in only 4/666 childcare programs, which was attributed to class distancing, the use of face masks for adults, universal symptom screening daily and disinfection.

### The impact of school closure on COVID-19 transmission

Eight published studies were identified with regard to the effect of school closure on the epidemiology of the COVID-19 pandemic, the results of which are presented in **Table 4**. Seven of these studies were based on mathematical modelling and various assumptions of infectivity from the first 3-4 months of the pandemic (23-29), while one was a time series study (30). The modelling studies in principal indicated that school closure is associated with a reduction in the number of cases, a reduction of hospitalizations and ICU admissions, with the effect of school closure dependent on the transmission rate, and the duration of school closure. Only one time series was reported by Auger et al. assessing data collected between 9 March 2020 and 7 May 2020, aiming at determining whether the closure of primary and secondary schools affects COVID-19 incidence and mortality in the USA (30). The results (adjusted for other enacted policies and testing rates) indicated that school closure was associated with a 62% reduction of COVID-19 incidence per week and a 58% reduction in mortality per week. It was found that countries which had a low cumulative COVID-19 incidence at the time of school closure, had greater reductions in incidence and mortality compared to those with a higher cumulative incidence at the same time.

**Table 4.**
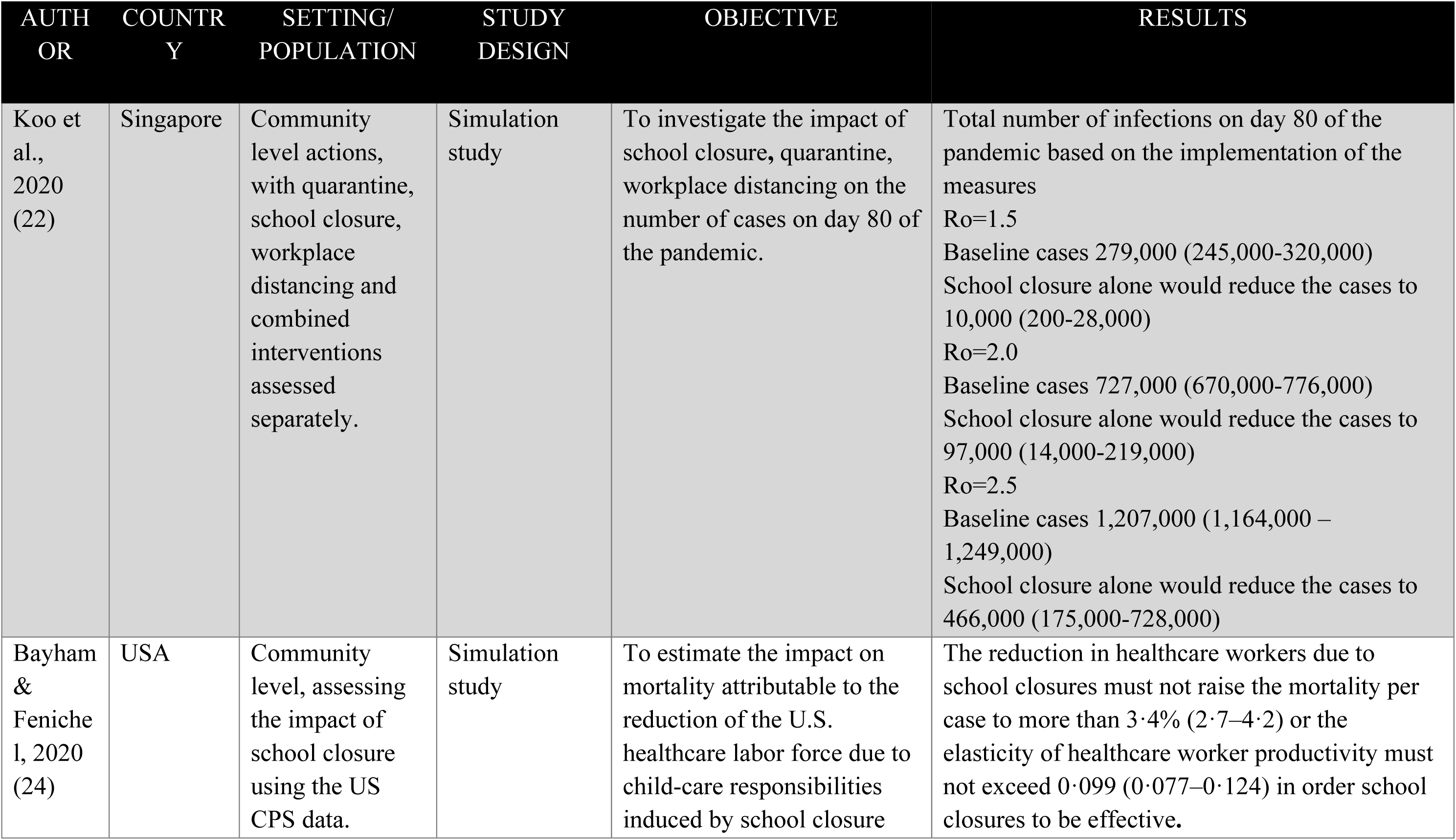
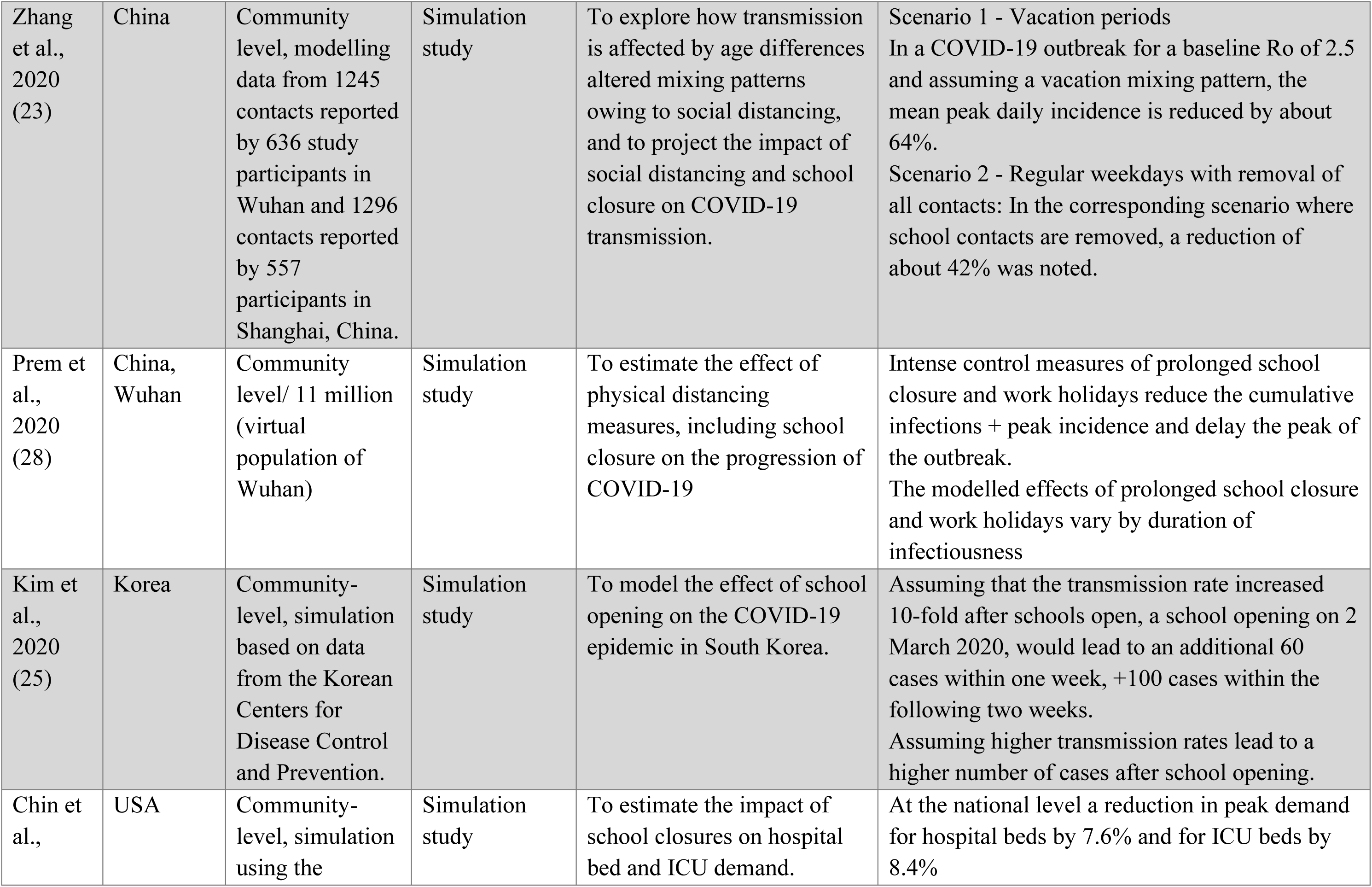
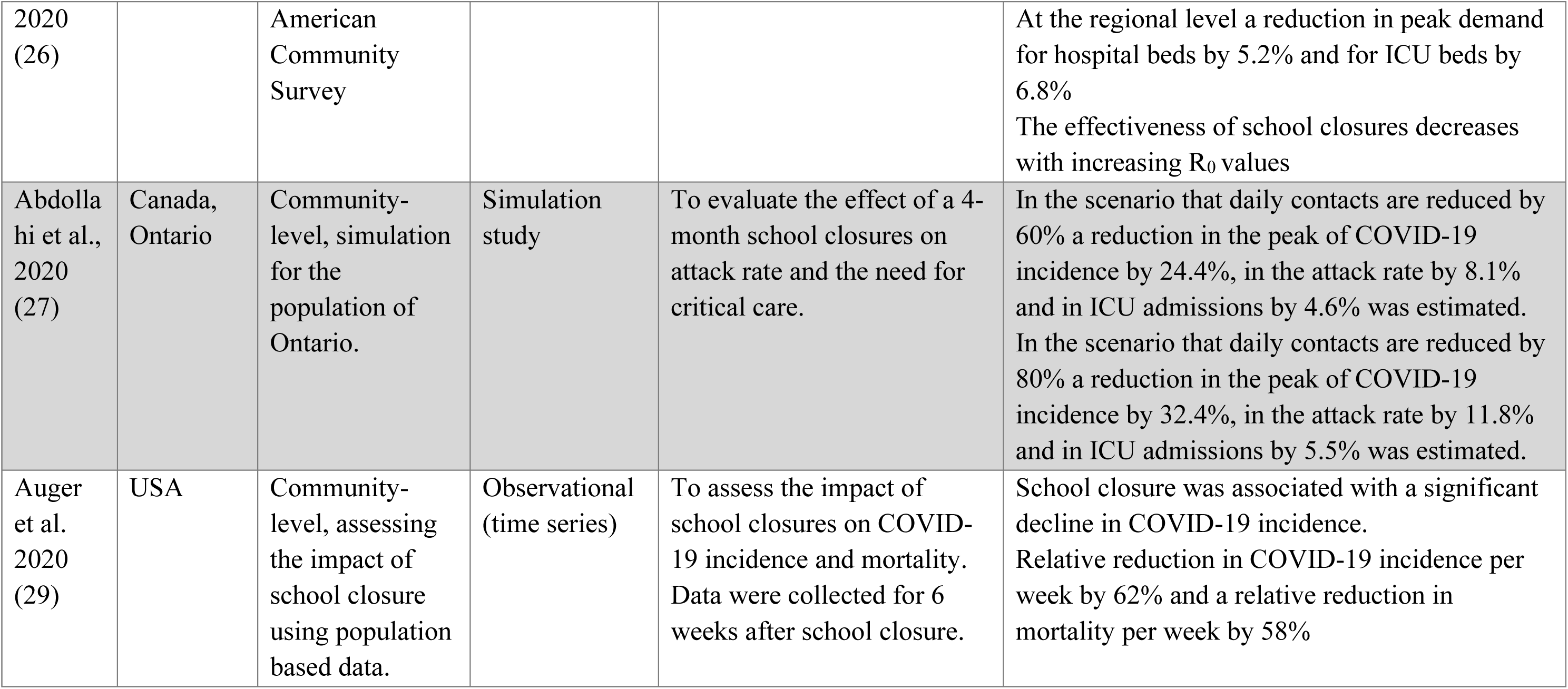
Results of the studies referring to the impact of school closures and SARS-CoV-2.

## Discussion

This study provides an up-to-date rapid review of the peer-reviewed literature pertaining to SARS-CoV-2 transmission by children, a topic which is a crucial input to assessments of the role of school settings in COVID-19 transmission (31). The literature appraised in this review provides sufficient evidence that children can both be infected by and transmit SARS-CoV-2 in both community, household and school settings. This finding is corroborated by research published after the cut-off date for this review, such as a large study from India (32).

Transmission of SARS-COV-2 has thus far been documented to be higher in household settings than in other community settings – including schools – a finding which may be potentially attributable to the individual, behavioural and contextual factors of households vs. other settings, as has been suggested elsewhere (33). Meanwhile, while children are overall noted to have lower rates of severe COVID-19 cases, there is evidence of differing transmission dynamics between younger vs. older children. In particular, there is some evidence that when index cases, younger children, such as under 10 years of age, lead to lower secondary attack rates than older children and adult. This matter warrants further investigation before concrete conclusions can be drawn (34, 35).

Within our review there were limited cases in the assessed studies in which a child index case was responsible for secondary transmission in schools, with the notable exception of an outbreak in Israel (21), which was associated with dense spacing, lack of the use of facemasks and closed spaces with poor ventilation. On the contrary, evidence from Rhode Island in the US, noted a very small number of cases after schools reopened during June-July 2020, which the authors attribute to the strict non-pharmaceutical interventions implemented including the use of face masks, physical distancing, screening for symptoms and classroom disinfection (22). A similar experience has been highlighted in Hong Kong, where multiple potential introductions of COVID-19 did not lead to secondary transmission, amidst a setting with numerous precautionary in-school measures, such as widespread facemask use, daily temperature checks, and staggered entries and exits (36).

In our review, seven modelling studies assessed the role of school closure, and overall indicated that school closure is associated with a reduction in the number of cases, hospitalisations and ICU admissions. However, the primary endpoints in the modelling studies were dependent on the transmission rate and the duration of school closure. While school closure may reduce SARS-CoV-2 transmission, the societal and economic impacts of prolonged school closure are noteworthy, as they may impact the availability of the healthcare workforce, (25, 37) and may also have negative effects on children through the interruption of the educational learning, social isolation, increased exposure to home violence, and rise in dropout rates (38).

There are important limitations to this study that may impact the direct implications for decision-making. As we assessed peer-reviewed evidence published in two biomedical databases, it inherently reflects the status quo of the end of the previous school year (2019- 2020) due to the lag time between study implementation, peer review and publication. A further limitation of this report refers to the study designs, notably case series and case reports. Finally, the household studies that we assessed reflect a broad geographical and temporal range are limited in comparability due to varying factors such as: background levels of community SARS-CoV-2 transmission; enrolment strategies; household structures and social isolation practices within households; study testing rates; and varying physical distancing policies including school closures.

Despite these limitations, the findings presented here provide an up-to-date assessment of the currently published peer-reviewed evidence. However, with an upsurge of cases in autumn 2020, the likelihood of introductions of SARS-CoV-2 into school settings increases. Continuous surveillance and assessment of the evidence is warranted to ensure the maximum protection of the health of students and the educational workforce. Where schools remain open, in-school mitigation measures, particularly for schools with older children, should be adopted and continually refined as new knowledge comes to light.

## Data Availability

This is a systematic review. All papers are referenced and searchable. ECDC data presented to complement the paper are available thorugh hyperlinks.

https://www.ecdc.europa.eu/en/covid-19/situation-updates

https://www.ecdc.europa.eu/en/publications-data/download-data-response-measures-covid-19

## Acknowledgments

We would like to thank Tommi Kärki and the ECDC team collecting data on COVID-19 response measures for Figure 2. We would like to thank Katerina Papathanasaki, Chrysa Chatzopoulou and Ioanna Lagou for their assistance in data archiving and report preparation. We acknowledge the bravery and dedication of educational professionals everywhere during these most challenging times. This work was financed by ECDC under direct service contract ECD.10660.

---

**Supplementary Table 1.** Search strategy.

| # | Searches | Results |
| --- | --- | --- |
| 1 | exp coronavirus/ | 14828 |
| 2 | exp coronavirus infections/ | 14705 |
| 3 | (Coronavir* or nCov or covid or Middle East Respiratory Syndrome or MERS or Severe Acute Respiratory Syndrome or SARS).ti,ab,tw. | 44118 |
| 4 | 1 or 2 or 3 | 50748 |
| 5 | (adolescent or (pre?school adj child) or child or infant or baby or toddler or juvenile).ti,ab,tw. | 830046 |
| 6 | (bab\$ or infant or child or boy or girl or teen\$ or school?child\$).ti,ab,tw. | 856745 |
| 7 | 5 or 6 | 1048284 |
| 8 | 4 and 7 | 604 |
| 9 | limit 8 to human | 515 |
| 10 | limit 9 to yr="2019 -Current" | 278 |
| 11 | limit 10 to english language | 263 |

Database(s): **Ovid MEDLINE(R) ALL** 1946 to June 22, 2020 Search Strategy:
| # | Searches | Results |
| --- | --- | --- |
| 1 | exp Coronavirus/ | 17477 |
| 2 | exp Coronavirus Infections/ | 16923 |
| 3 | (Coronavir* or nCov or covid or covid-19 or Middle East Respiratory Syndrome or MERS or Severe Acute Respiratory Syndrome or SARS).ti,ab,kf. | 46333 |
| 4 | 1 or 2 or 3 | 51033 |
| 5 | (baby or babies or infant* or child* or boy* or girl* or toddler* or preschool* or pre?school* or teen* or schoolchild* or adolescen* or juvenil*).tw. | 2026780 |
| 6 | 4 and 5 | 2184 |
| 7 | humans.sh. | 18540514 |
| 8 | 6 and 7 | 1161 |
| 9 | limit 8 to yr="2019 -Current" | 340 |
| 10 | limit 9 to english language | 295 |

## References

1. ECDC. Download data on country response measures to COVID-19 2020 [Available from: https://www.ecdc.europa.eu/en/publications-data/download-data-response-measures-covid-19.

2. Viner RM, Mytton OT, Bonell C, Melendez-Torres GJ, Ward J, Hudson L, et al. Susceptibility to SARS-CoV-2 Infection Among Children and Adolescents Compared With Adults: A Systematic Review and Meta-analysis. JAMA Pediatr. 2020.

3. Jones TC, Mühlemann B, Veith T, Biele G, Zuchowski M, Hoffmann J, et al. An analysis of SARS-CoV-2 viral load by patient age. medRxiv. 2020:2020.06.08.20125484.

4. Heald-Sargent T, Muller WJ, Zheng X, Rippe J, Patel AB, Kociolek LK. Age-related differences in nasopharyngeal severe acute respiratory syndrome coronavirus 2 (SARS-CoV-2) levels in patients with mild to moderate coronavirus disease 2019 (COVID-19). JAMA Pediatr. 2020;174(9):902–3.

5. Götzinger F, Santiago-García B, Noguera-Julián A, Lanaspa M, Lancella L, Calò Carducci FI, et al. COVID-19 in children and adolescents in Europe: a multinational, multicentre cohort study. Lancet Child Adolesc Health. 2020;4(9):653–61.

6. Dong Y, Dong Y, Mo X, Hu Y, Qi X, Jiang F, et al. Epidemiology of COVID-19 among children in China. Pediatrics. 2020;145(6).

7. Milani GP, Bottino I, Rocchi A, Marchisio P, Elli S, Agostoni C, et al. Frequency of Children vs Adults Carrying Severe Acute Respiratory Syndrome Coronavirus 2 Asymptomatically. JAMA Pediatr. 2020.

8. Moher D, Liberati A, Tetzlaff J, Altman DG, Altman D, Antes G, et al. Preferred reporting items for systematic reviews and meta-analyses: The PRISMA statement. PLoS Med. 2009;6(7).

9. Investigation of a nosocomial outbreak of coronavirus disease 2019 in a paediatric ward in South Korea: successful control by early detection and extensive contact tracing with testing, (2020).

10. Goldstein E, Lipsitch M. Temporal rise in the proportion of younger adults and older adolescents among coronavirus disease (COVID-19) cases following the introduction of physical distancing measures, Germany, March to April 2020. Eurosurveillance. 2020;25:2000596.

11. Posfay-Barbe KM, Wagner N, Gauthey M, Moussaoui D, Loevy N, Diana A, et al. COVID-19 in Children and the Dynamics of Infection in Families. Pediatrics. 2020;146:e20201576.

12. Teherani MF, Kao CM, Camacho-Gonzalez A, Banskota S, Shane AL, Linam WM, et al. Burden of illness in households with SARS-CoV-2 infected children. Journal of the Pediatric Infectious Diseases Society. 2020.

13. Somekh E, Gleyzer A, Heller E, Lopian M, Kashani-Ligumski L, Czeiger S, et al. The Role of Children in the Dynamics of Intra Family Coronavirus 2019 Spread in Densely Populated Area. The Pediatric Infectious Disease Journal. 2020;39(8).

14. Maltezou HC, Vorou R, Papadima K, Kossyvakis A, Spanakis N, Gioula G, et al. Transmission dynamics of SARS-CoV-2 within families with children in Greece: A study of 23 clusters. Journal of Medical Virology. 2020:0–3.

15. Park YJ, Choe YJ, Park O, Park SY, Kim Y-M, Kim J, et al. Contact Tracing during Coronavirus Disease Outbreak, South Korea, 2020. Emerging Infectious Disease journal. 2020;26(10):2465.

16. Kim J, Choe YJ, Lee J, Park YJ, Park O, Han MS, et al. Role of children in household transmission of COVID-19. Archives of Disease in Childhood. 2020:1–3.

17. Heavey L, Casey G, Kelly C, Kelly D, McDarby G. No evidence of secondary transmission of COVID-19 from children attending school in Ireland, 2020. Eurosurveillance. 2020;25:2000903.

18. Danis K, Epaulard O, Bénet T, Gaymard A, Campoy S, Botelho-Nevers E, et al. Cluster of Coronavirus Disease 2019 (COVID-19) in the French Alps, February 2020. Clinical Infectious Diseases. 2020;71(15):825–32.

19. Yung CF, Kam K-q, Nadua KD, Chong CY, Tan NWH, Li J, et al. Novel coronavirus 2019 transmission risk in educational settings. Clinical Infectious Diseases. 2020:3–6.

20. Macartney K, Quinn HE, Pillsbury AJ, Koirala A, Deng L, Winkler N, et al. Transmission of SARS-CoV-2 in Australian educational settings: a prospective cohort study. Lancet Child Adolesc Health. 2020;4642:1–10.

21. Stein-Zamir C, Abramson N, Shoob H, Libal E, Bitan M, Cardash T, et al. A large COVID-19 outbreak in a high school 10 days after schools’ reopening, Israel, May 2020. Eurosurveillance. 2020;25:1–5.

22. Link-Gelles R, DellaGrotta AL, Molina C, Clyne A, Campagna K, Lanzieri TM, et al. Limited Secondary Transmission of SARS-CoV-2 in Child Care Programs — Rhode Island, June 1–July 31, 2020. MMWR Morbidity and Mortality Weekly Report. 2020;69:1170–2.

23. Koo JR, Cook AR, Park M, Sun Y, Sun H, Lim JT, et al. Interventions to mitigate early spread of SARS-CoV-2 in Singapore: a modelling study. The Lancet Infectious Diseases. 2020;20:678–88.

24. Zhang J, Litvinova M, Liang Y, Wang Y, Wang W, Zhao S, et al. Changes in contact patterns shape the dynamics of the COVID-19 outbreak in China. Science (New York, NY). 2020;368:1481–6.

25. Bayham J, Fenichel EP. Impact of school closures for COVID-19 on the US health-care workforce and net mortality: a modelling study. The Lancet Public Health. 2020;5:e271–e8.

26. Kim S, Kim YJ, Peck KR, Jung E. School opening delay effect on transmission dynamics of Coronavirus disease 2019 in Korea: Based on mathematical modeling and simulation study. Journal of Korean Medical Science. 2020;35.

27. Chin ET, Huynh BQ, Lo NC, Hastie T, Basu S. Projected geographic disparities in healthcare worker absenteeism from COVID-19 school closures and the economic feasibility of child care subsidies: a simulation study. BMC medicine. 2020;18:218.

28. Abdollahi E, Haworth-Brockman M, Keynan Y, Langley JM, Moghadas SM. Simulating the effect of school closure during COVID-19 outbreaks in Ontario, Canada. BMC Medicine. 2020;18:1–8.

29. Prem K, Liu Y, Russell TW, Kucharski AJ, Eggo RM, Davies N, et al. The effect of control strategies to reduce social mixing on outcomes of the COVID-19 epidemic in Wuhan, China: a modelling study. The Lancet Public Health. 2020;5:e261–e70.

30. Auger KA, Shah SS, Richardson T, Hartley D, Hall M, Warniment A, et al. Association between Statewide School Closure and COVID-19 Incidence and Mortality in the US. JAMA - Journal of the American Medical Association. 2020;45229:859–70.

31. ECDC. COVID-19 in children and the role of school settings in COVID-19 transmission. Solna, Sweden: European Centre for Disease Prevention and Control; 2020 August 6.

32. Laxminarayan R, Wahl B, Dudala SR, Gopal K, Mohan C, Neelima S, et al. Epidemiology and transmission dynamics of COVID-19 in two Indian states. Science. 2020:eabd7672.

33. Merckx J, Labrecque JA, Kaufman JS. Transmission of SARS-CoV-2 by Children. Dtsch Arztebl International. 2020;117(33-34):553–60.

34. Cho EY, Choi EH, Kim J-H. Interpreting Transmissibility of COVID-19 in Children. Emerging Infectious Disease journal. 2020;26(12).

35. Goldstein E, Lipsitch M, Cevik M. On the effect of age on the transmission of SARS-CoV-2 in households, schools and the community. medRxiv. 2020:2020.07.19.20157362.

36. Fong M, Cowling B, Leung G, Wu P. Letter to the editor: COVID-19 cases among school-aged children and school-based measures in Hong Kong, July 2020. Eurosurveillance. 2020;25(37):2001671.

37. Sadique MZ, Adams EJ, Edmunds WJ. Estimating the costs of school closure for mitigating an influenza pandemic. BMC Public Health. 2008;8(1):135.

38. UNESCO. Adverse consequences of school closures. Unesco. 2020.

